# Dose-finding, experimental medicine evaluation of sodium valproate for the prevention of post-cardiac surgery myocardial injury

**DOI:** 10.64898/2026.08.30.26361746

**Authors:** Marius Roman, Natasha Beasley, Shameem Ladak, Charles Solomon, Weiqi Laio, Florence Y Lai, Lathishia Joel-David, Hardeep Aujla, Gianluigi Condorelli, Marcin J Woźniak, Veryan Codd, Tom Webb, Cassey Brookes, Hussein Mulla, Gerry P McCann, Gavin J Murphy

## Abstract

**Background:** A dose-finding trial evaluated safety and adherence for pre-cardiac surgery administration of sodium valproate. Integrated multi-omics analyses of myocardium were used to characterise mechanisms underlying treatment effects.

**Methods:** Adults undergoing cardiac surgery were randomised 1:1:1:1 with concealed allocation to no treatment (Controls), sodium valproate 15mg/kg/day for 1-2 weeks, 15mg/kg/day for 4-6 weeks, or 25mg/kg/day for 4-6 weeks pre-surgery. The primary analysis evaluated adherence and toxicity. Myocardial injury was defined by high sensitivity serum troponin at 24 hours post-surgery. Single-nucleus Assay for Transposase-Accessible Chromatin with sequencing (snATACseq) and single nuclei RNA sequencing (snRNAseq) of myocardial biopsies collected at surgery assessed treatment effects on chromatin accessibility and gene expression. Candidate mechanisms were validated in *in vitro*.

**Results:** The analysis cohort included 42 participants enrolled between January 2020 and August 2024. Sodium valproate 15mg/kg/day for 1-2 weeks had the highest levels of complete treatment adherence (70%), with 20% experiencing moderate/severe drug related adverse effects. Non-compliance was increased with longer and higher dosing.

An as-treated analyses demonstrated reductions in troponin release in participants receiving Valproate≤14 days where myocardial biopsies demonstrated activation of hormetic p53 and Akt-GSK-3β ferroptosis protection pathways versus controls. Silencing of these pathways attenuated the protective effect of valproate *in vitro*. Treatment effects were not attributable to chromatin accessibility.

Treatment >14 days resulted in a heart failure phenotype, suppression of ferroptosis protection pathways, and increased myocardial injury.

**Conclusions:** Valproate 15mg/kg/day for ≤14 days pre-surgery is well tolerated in adults awaiting cardiac surgery and is associated with upregulation of ferroptosis protection pathways and reductions in myocardial injury.

## Introduction

Perioperative myocardial injury affects >25% of cardiac surgery procedures where it is the primary cause of early death, or results in delayed recovery, lower quality of life, and increased healthcare resource use.(1) Decades of research have not delivered effective myocardial protection interventions.(2–6)

Sodium valproate is a class I/IIa Histone Deacetylase inhibitor (HDACi) in widespread use for the treatment of neurological diseases that protects myocardium from acute metabolic stress in a wide range of pre-clinical models.(7) We hypothesised that pre-surgery administration of sodium valproate would have myocardial protection effects in people exposed to the severe metabolic stress of cardiac surgery.

Valproate has unpredictable pharmacodynamics where *in vivo* effectiveness is not directly attributable to plasma concentrations. Valproate also has both predictable (GI symptoms, reduction in platelet counts) and unpredictable (hepatotoxicity, pancreatitis) toxicity that affect adherence and safety. The current trial was designed as a dose finding study with pre-specified adherence, safety, and pharmacodynamic progression criteria to an effectiveness evaluation. Based on the known biological effects of valproate, in an experimental sub-study we further investigated whether treatment effects would be attributable to known effects of valproate on chromatin accessibility (8) and pro-survival gene expression.(7–9)

## Methods

### Trial Design

A Randomised Controlled Trial of Pre-Surgery Sodium Valproate for the Prevention of Organ Injury in Cardiac Surgery (Val-CARD) was an open label single blinded parallel group RCT conducted at a single tertiary cardiac surgery centre in the UK. The protocol was registered prospectively (NCT03825250). All participants had provided informed consent, and ethical approval was obtained prospectively from the East Midlands Research Ethics Committee (18/EM/0188). Recruitment was suspended between March 2020 and September 2022 due to COVID-19. Trail recruitment was terminated after the completion of the dose finding study in September 2024 due to exhaustion of funds.

### Participants

Adult cardiac surgery patients (≥18 years) undergoing cardiac surgery (CABG, Valve, or CABG and Valve) able in the opinion of the investigator, and willing to give informed consent were eligible. Patients undergoing urgent, emergency or salvage procedures, or those with Stage 4 CKD, chronic atrial fibrillation, a platelet count ≤150×10^9^.ml^-1^, liver disease, or a recognised contra-indication to sodium valproate were ineligible.

### Randomisation

Participants were randomised with concealed allocation using Sealed Envelope in a 1:1:1:1 ratio to:

1. Standard care (no treatment)
2. Sodium valproate at a target dose of 15mg/kg per day for 1-2 weeks pre-surgery.
3. Sodium valproate at a target dose of 15 mg/kg per day for 4-6 weeks pre-surgery.
4. Sodium valproate at a target dose of 25 mg/kg per day for 4-6 weeks pre-surgery.

### Intervention

Sodium Valproate was administered as a twice daily oral dose for 1-6 weeks pre-surgery. To minimise toxicity all participants commenced at 10mg/kg/day with dose escalation by 2.5mg/kg/day (max 200mg) every three days to achieve the target dose as specified in the treatment allocations.

Flexibility in target treatment duration anticipated up to 30% of elective cardiac surgery procedures being rescheduled at short notice to reduce protocol non-compliance.

Participants in the control arm received standard care; no treatment.

### Monitoring

Safety and toxicity evaluations included baseline measurement of blood counts with assessment of hepatic and renal function, plasma valproate levels and full blood counts at baseline and after every two weeks of treatment. Participants underwent twice weekly telephone call-based monitoring during the intervention period to assess compliance and adverse effects, and to advise on dose escalation. Dose escalation was based on assessments of tolerance and likely adverse effect as follows:

1. Patients experiencing no adverse effects were advised to continue dose escalation.
2. Patients experiencing no adverse events were advised to continue escalation.
3. Patients with Class I Common Terminology Criteria for Adverse Events (CTCAE, https://evs.nci.nih.gov/ftp1/CTCAE/About.html) adverse events were advised to continue at the current dose if tolerated or to reduce the dose if not.
4. Patients experiencing Grade 2 CTCAE were advised to reduce the dose to that previously tolerated.
5. Patients with Grade 3-5 CTCAE were advised to stop the treatment.

### Outcomes

Prespecified primary outcomes for Phase 1 were:

1. Protocol compliance.
2. Accumulated valproate dose.
3. Trough plasma valproate concentrations measured at 1-2 weeks post commencement (Group D), and at 4 and 6 weeks (Groups A, B and C).

A pre-specified secondary analysis explored pharmacodynamics, specifically dose-responses versus biomarkers of myocardial and kidney injury estimated from Area Under the Curve from baseline to 96 hours postoperatively for high sensitivity serum Troponin I (hsTroponin I) concentrations (Enzo® Troponin I (human) ELISA kit, Enzo Life Sciences, Ann Arbor, MI, USA) and serum creatinine. Adverse events were as defined by CTCAE, other than those included in the primary and secondary endpoints. Myocardial injury was as defined by Devereaux and colleagues.(10)

### Statistical Considerations

A sample size of 40 participants was considered adequate to assess compliance and toxicity based on UK guidance for feasibility studies (11) and our previous experience of dose finding studies (12) in the target population.

The intention to treat analysis included all trial participants who underwent surgery. Baseline characteristics were presented by descriptive statistics, using mean and standard deviation (SD) for continuous data, and frequency (N) and percentage (%) for binary and categorical data in the full analysis set (FAS). One-way analysis of variance (ANOVA) and Pearson’s chi-square χ2 test were used to investigate the differences in baseline characteristics among the four groups for continuous and binary/categorical variables, respectively. Safety and toxicity endpoints (thrombocytopenia and hepatotoxicity) were analysed in all participants who at least took one dose of the randomised treatment (safety population set, SAF) and summarised by descriptive statistics. Generalized Estimating Equations (GEE) were used to explore the population average of the treatment effect (group effect), time effect, and the interaction between treatment and time effects on the two primary outcomes (troponin and serum creatinine) measured across several time points from baseline to 6 weeks after cardiac surgery. Compared with the baseline measurements, the changes in troponin and serum creatinine over time were presented in box and whisker plots and line charts by randomised groups and time points. All statistical analyses were performed in Stata 18.

Due to high levels of non-adherence to valproate treatment the mechanism sub study used an as-treated approach that was not pre-specified in the trial protocol. As non-adherence with higher doses and longer treatment resulted in comparable drug exposure in the valproate 15mg/day for 4-6 weeks and the 25mg/kg/day for 4-6 weeks groups, these were considered as a single group in the as-treated analyses.

### Transcriptomics and Chromatin Accessibility

snRNAseq, was performed as described.(13) cDNA amplification and library construction were performed using Chromium Single Cell 3’ Gel-bead in Emulsion (GEM) and Chromium Single Cell 3’ Library kits v3.1. (#1000128, 10x Genomics, Pleasanton, United States) following the manufacturer’s instructions. Briefly, cells were re-suspended in the required concentration for the desired targeted cell recovery and combined with a reverse transcription master mix. This was loaded onto the Chromium B Chip and the instrument run. The post GEM/reverse-transcribed product was collected, and cDNA amplification performed. The resulting cDNA quality-checked using the Agilent Bioanalyzer High Sensitivity DNA kit (Agilent Technologies, Santa Clara, United States). Libraries were constructed, including fragmentation, end-repair, A-tailing, and adaptor ligation steps. A unique sample index was added to each sample. Following library construction, samples were quality-checked, and libraries was purified and eluted using Buffer EB (#19086, Qiagen, Hilden, Germany), and SPRIselect beads (#B23317, Beckman Coulter, Pasadena, United States). Samples was sequenced paired-end on Illumina NovoSeq at 2.5×108 reads per sample and analysed as described previously.(14)

For single-nuclei ATAC sequencing, nuclei isolation and library construction used the Chromium Next GEM Single Cell ATAC Library kit v1.1. (10x Genomics). Briefly, nuclei were re-suspended at the required concentration for the desired targeted nuclei recovery and transposed using a transposition mix. Transposed nuclei were loaded on Chromium H Chip and the instrument run. After library construction, a unique sample index was added to each sample. Following quality control, libraries were purified and eluted using Buffer EB and SPRI select beads, with sequencing as described above, and bioinformatic analyses as described below.

### Single-nuclei ATAC-sequencing Data Processing and Quality Control

Raw multiome sequencing data were processed using Cell Ranger ARC v2.1.0 (10x Genomics), with FASTQ files aligned to the human reference genome (GRCh38-2024-A) using default parameters. Fragment files, peak accessibility matrices and associated quality metrics were generated for each sample.

Chromatin accessibility data was analysed in R (v4.4.2) using the Signac (15) and Seurat (16) packages. Peak accessibility matrices were imported from the CellRanger ARC filtered feature-barcode matrices and used to generate ChromatinAssay objects, with gene annotations were obtained from Ensembl release 86 (EnsDb.Hsapiens.v86) and converted to hg38 coordinates (17). Quality control metrics, including the TSS enrichment and nucleosome signal scores, were calculated for all nuclei. Nuclei were retained if they met the following criteria; ATAC fragment count >800 and <100,000, nucleosome signal <1, TSS enrichment >1. Samples yielding fewer than 50 nuclei post-filtering were excluded.

Filtered datasets were merged and chromatin accessibility data normalised using TF-IDF transformation. Dimensionality reduction was performed by LSI via SVD of the TF-IDF matrix, and UMAP was applied to LSI dimensions 2-30 for visualisation. Cell type annotation was performed using a reference-based approach, using a human cardiac reference atlas (SCP1849; ICM_scportal_05.24.2022.h5ad), with transfer anchors identified and cell identities predicted using Seurats FindTransferAnchors function and MapQuery functions. Predicted labels were used for all downstream chromatin accessibility analyses. Ambient RNA contamination was removed using DecontX with decontaminated counts retained for expression-based analyses (18).

### Chromatin and Promoter accessibility analysis

Cardiomyocytes and endothelial cells were analysed separately according to transferred cell-type labels. To minimise biases arising from unequal cell numbers, each pairwise comparison was balanced by random down sampling to the size of the smallest group prior to differential accessibility testing.

Peaks were annotated using ChIPseeker with the TxDb.Hsapiens.UCSC.hg38.knownGene annotation database (19). Promoter accessibility analyses were performed by overlapping ATAC peaks with regions defined as ±2kbp around annotated TSS, with peaks assigned to genes by coordinate overlap. Where multiple promoter peaks were mapped to the same gene, the peak with the largest accessibility peak was retained. Differentially accessible regions (DARs) were identified using logistic regression via Seurats FindMarkers, with total ATAC fragment counts included as a covariate and sex included where it varied between groups. Significance thresholds of FDR<0.05 and log2FC≥0.25 were applied throughout.

Concordance between promoter accessibility and gene expression was assessed by gene-level fold across modalities using Spearman ranks correlation, with genes classified as exhibiting concordant or discordant relationships. TF motif enrichment was performed using Signac with JASPAR2020 position frequency matrices (15). Promoter-associated DARs were stratified by direction of change tested against the full ATAC peak set as background using FindMotifs, with enriched motifs ranked by fold enrichment and adjusted p-value.

### Single-nuclei RNA-sequencing Data Processing and Quality Control

Gene expression matrices from Cell Ranger ARC v2.1.0 were quality-controlled independently per sample, retaining nuclei with 200-12,500 detected genes and less than 5% mitochondrial transcript content. Doublets were identified and removed using DoubletFinder (20) following SCTransform and PCA. Optimal pK values were determined using parameter sweeps, with the expected doublet rate was set to 0.8% per sample. Samples containing fewer than 50 nuclei post-QC were excluded. Remaining samples were merged into a single Seurat object and cell-type labels assigned using the same reference atlas and MapQuery label transfer approach as described above.

### Differential Gene Expression Analysis

Differential expression testing was performed using logistic regression via Seurat FindMarkers function, requiring genes to be detected in ≥10% of nuclei in either group, with log2FC≥0.25. Decontaminated transcript counts were included as a latent variable, and sex was included as an additional covariate where applicable. Significance was defined as FDR<0.05 (Benjamini-Hochberg correction) and log2FC≥0.25.

Functional enrichment analyses of significantly up- and downregulated genes was performed using ClusterProfiler (21) for Gene Ontology (GO) Biological Processes (22) and KEGG (23) pathways using all tested genes as the background universe. Enriched pathways were considered significant at an adjusted p-value <0.05.

TF activity was inferred using DoRothEA regulon database (24) (confidence levels A-C) and the VIPER algorithm (25), with differential activity assessed by logistic regression as above. To distinguish regulatory from transcriptional changes, differential expression was also performed on TF-encoded genes, and results integrated across analyses. TFs with concordant directional changes in both activity and expression were classified as concordant, and those with opposing directions as discordant.

Cell-cell communication was analysed using CellChat (26) with the CellChatDB human ligand-receptor database. Communication probabilities were calculated using a trimmed mean (trim=0.1), excluding interactions supported by fewer than 10 cells. Pathway-level signalling differences between treatment groups were assessed by comparing information flow and ranked by changes in aggregated communication probability.

### In vitro experiments

The cardioprotective effects of valproate were evaluated in AC16 cardiomyocytes pre-stressed with IL-6 + H_2_O_2_ *in vitro*. These demonstrate increased susceptibility to IRI as well as transcriptional overlap with cardiomyocytes from myocardial injury susceptibility injury patients undergoing cardiac surgery.(14)

AC16 human cardiomyocyte cell lines were maintained at 37 °C in 5% CO2 in DMEM: F12 medium (Gibco, 11580546) supplemented with 12.5 % FBS and (27) Simulation of Cluster 1 used 4 hrs exposure to H_2_0_2_ 100µM and IL6 10ng.ml^-1^ to model the ROS and IL-6 signalling observed in patients demonstrating myocardial injury susceptibility *in vivo.*(14) Cells cultured only in total medium served as controls. Next Generation bulk mRNAseq (Source Genomic Ltd, Nottingham UK) of AC16 cells was used to estimate homology to the snRNAseq analysis. Based on the transcriptomic data we tested whether myocardial protection by valproate was attributable to hormetic p53 activation via hyperacetylation, and PI3K/AKT/GSK3β activation. Gene silencing was achieved by transfection with Lipofectamine RNAiMAX (Invitrogen, Carlsbad, CA, USA, 13778075) transfection reagent at 5–50 nmol/L final concentration of p53 siRNA (ThermoFisher Scientific, Silencer Select; Assay ID: 106141), AKT1 siRNA (ThermoFisher Scientific, Silencer Select; Assay ID: 42811), GSK3β siRNA (ThermoFisher Scientific, Silencer Select; Assay ID: 42839), or

Silencer™ Select Negative Control No. 1 siRNA (ThermoFisher Scientific, 4390843) in Opti-MEM reduced serum media (ThermoFisher Scientific, 31985070) according to the manufacturer’s protocol for 24 hours.

Ischaemia reperfusion injury (IRI) was simulated by culture in glucose deficient media in anoxic conditions (95% N_2_ and 5% CO_2_) at 37°C for 4 hrs, followed by replacing glucose free media with total medium at 37°C in a 95% air and 5% CO_2_ (normoxic condition) incubator for 24 hrs. Cells cultured only in normoxic conditions served as controls. Cardiomyocyte injury was assessed using the LDH-Cytox™ Assay Kit (Biolegend, 426401). All experiments used 3-4 repeats.

Total RNA from AC16 cells was isolated using RNeasy kit (Qiagen, Venlo, The Netherlands, 74104) and cDNA was synthesised using Tetro cDNA synthesis kit (Meridian Bioscience, Cincinnati, OH, USA, BIO-65043). Gene expression was quantified by quantitative real time PCR using TaqMan primers (ThermoFisher Scientific, Assay IDs: Hs01005664_m1 and Hs00608023_m1) and SensiFast Probe Hi-ROX kit (Meridian Bioscience, Cincinnati, OH, USA, BIO-82005) on Rotor gene Q (Qiagen) using the manufacturer’s protocol. Relative levels were calculated using the 2−(ΔΔCt) method, and mRNA expression was normalised to housekeeping gene PPIA (ThermoFisher Scientific, Assay ID: Hs99999904_m1).

## Results

### Trial cohort

There were 45 participants recruited and randomised between September 2019 and September 2024 at a single UK centre. Of these 3 were withdrawn prior to surgery leaving an analysis population of 42 participants. Follow-up was 100% complete. No participants died during the trial. Details of participant flow are listed in **Figure 1**. Baseline demographic, clinical, and operative characteristics were well balanced across all 4 groups (**eTable 1**).

**Figure 1.**
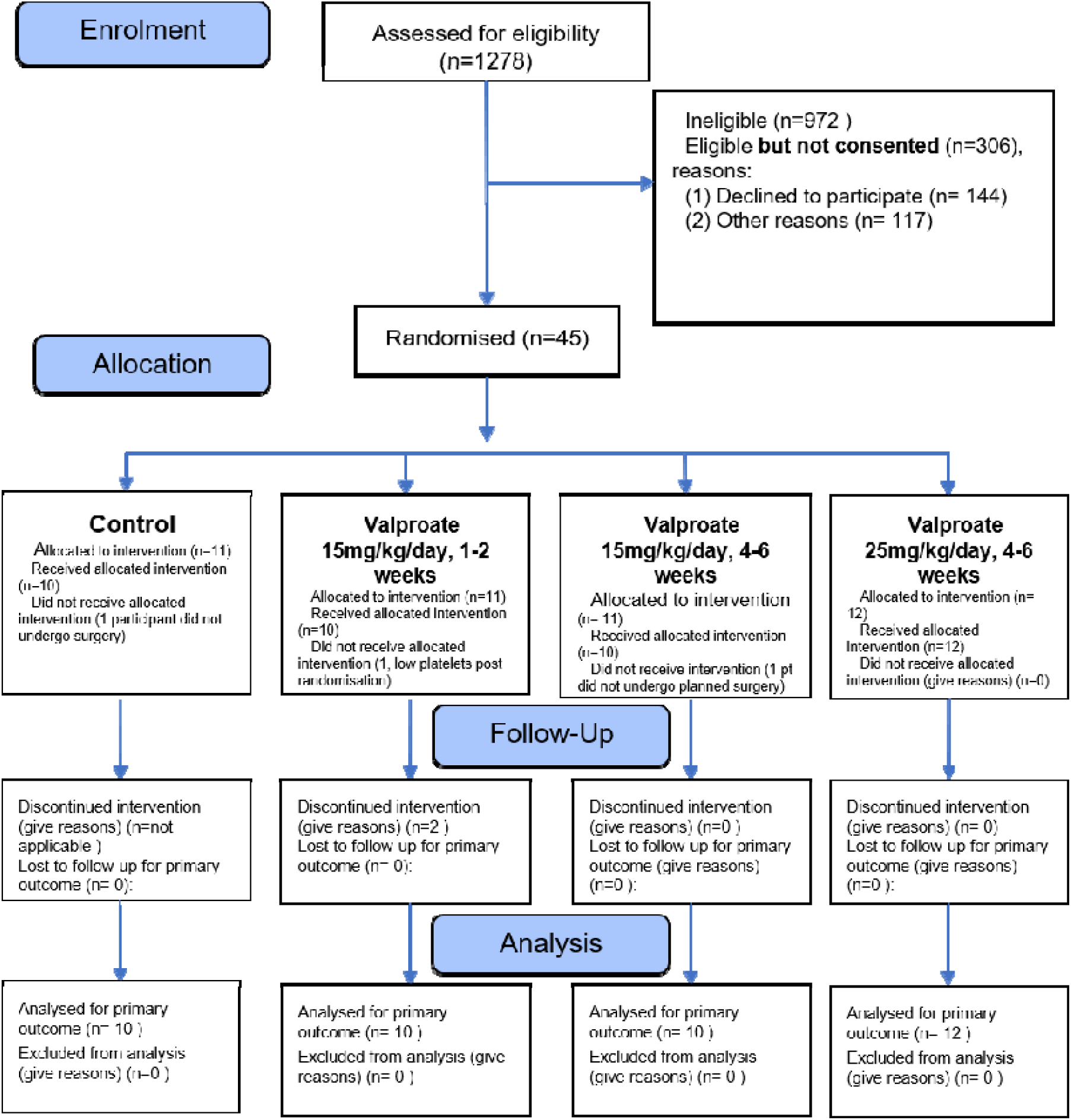
Intention to treat analyses. CONSORT diagram of participant flow in the VALCARD dose finding analysis.

### Intention to Treat Analysis

Adherence to allocated treatment and adverse events are shown in **Table 1** and **eTable 2**. Patients allocated to Valproate 15mg/kg/day for 1-2 weeks, had the highest level of complete adherence to protocol; 70%, with 90% of participants reaching their target dose and 70% of participants received all allocated doses. The Valproate 15mg/kg/day for 1-2 weeks group reported CTCAE events with severity ≥2 in 2 participants, and the lowest rates of new platelet count ≤150×10^9^.ml^-1^; 56% versus other groups. Rates of abnormal liver function tests were low (≤10%) and similar between groups (**eTable 2**). There was no treatment interaction for group allocation and serial measures of hsTroponin and creatinine from baseline to 96 hours (**eTable 3**).

**Table 1.**
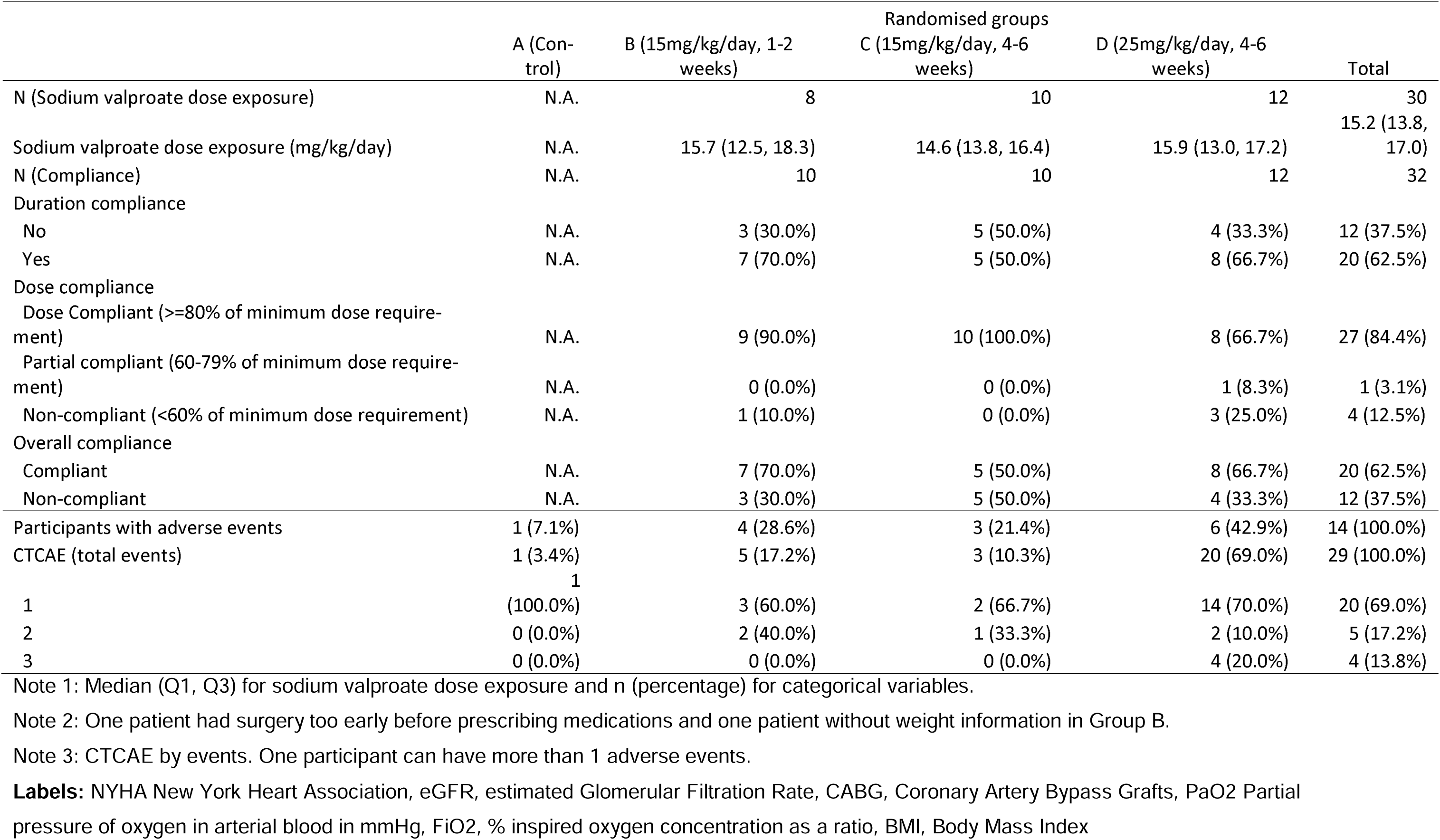
Treatment compliance, protocol deviations and adverse events in the Intention to Treat Analysis.

### As treated analyses

In the as treated analysis 11 participants were allocated to **Group A** (no Valproate), 12 to **Group B** (Valproate≤14 days), and 19 to **Group C** (Valproate>14 days). Demographic, clinical, and operative characteristics for the as-treated analyses were not different between groups (**eTable 4)**. Area under the curve for hsTroponin I from surgery to 96 hours for individual trial participants by treatment duration are shown in **Figure 2**. A dose response curve fitted using fractional polynomials suggested that the nadir troponin release occurs between 5-10 days of treatment. In participants who underwent surgery with cardioplegic arrest (IRI) the frequency of myocardial injury was 44% (n=4/9) in Controls, 22% (n= 2/9) in Valproate≤14 days and 73% (n=11/15) in Valproate>14 days, Chi Squared p=0.047 (**eTable 4**).

**Figure 2.**
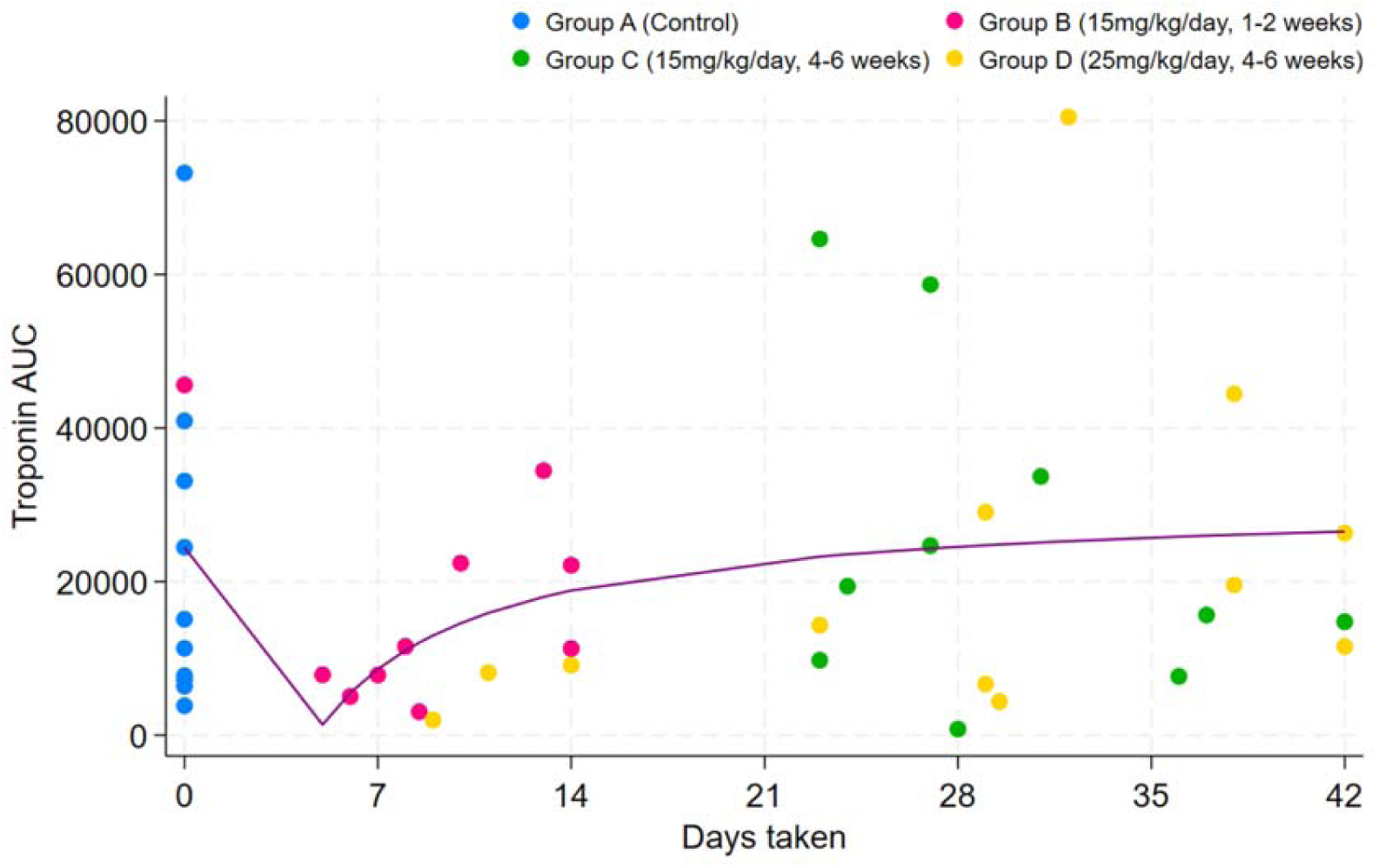
Area under the curve for serial troponin measurements from surgery to 96 hours post-surgery for trial participants. Colours represent treatment allocation. The dose response curve was fitted using fractional polynomials.

### Transcriptomics Analyses

snRNAseq and snATACseq analyses (n=18 biopsies) focused on cardiomyocytes (1803 nuclei), and endothelial cells (6,006 nuclei) as key determinants of the myocardial response to IRI.

#### Cardiomyocytes

*For Valproate*≤*14 days versus Controls*, snATACseq demonstrated a large net increase in significant Differentially Accessible Regions (DAR, 37 up versus 11 down), including gene promoters, introns, and distal intergenic regions (**Figure 3A, eTable 5-7**). There was complete discordance between DAR and Differentially Expressed Genes (DEG) shown with snRNAseq (**Figure 3B-D**).

**Figure 3.**
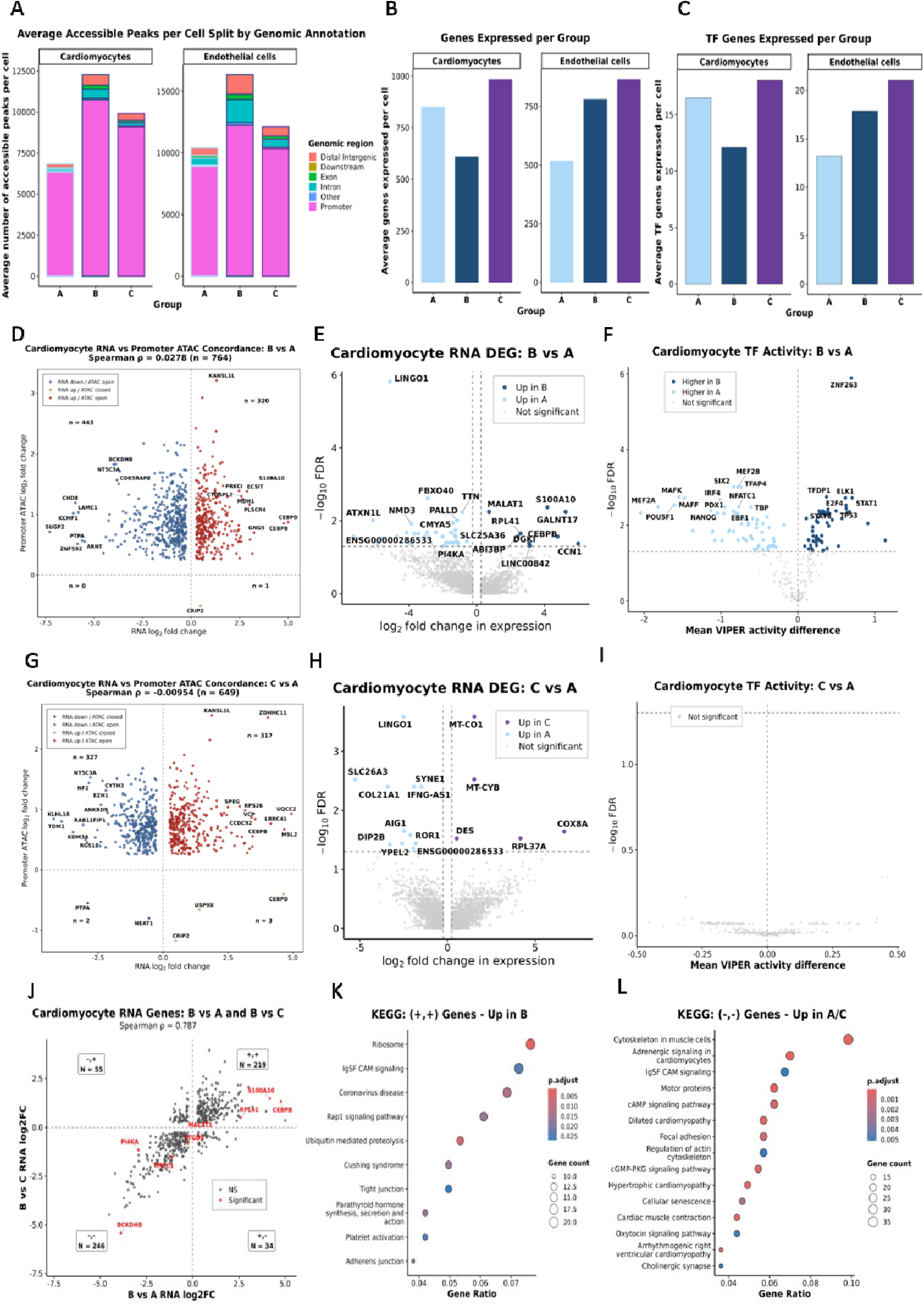
As treated comparison of chromatin accessibility, gene expression, and transcription factor activity in cardiomyocytes. **A.** Chromatin accessibility shown as average accessible peaks per cell split by genomic annotation in snATACseq analyses. **B.** Total number of differentially expressed genes by cell type versus as-treated group. **C.** Differential Transcription factor activation shown as differentially expressed TF target genes versus as treated group. **D.** Quadrant plot of Differential Accessible Regions (DAR) versus Differentially Expressed Genes for the Valproate≤14 days (Group B) versus Controls (Group A) comparisons. Volcano plots showing top 20 **E.** DEG and **F.** Differential TF activation in Cardiomyocytes for Valproate≤14 days (Group B) versus Controls (Group A) comparisons. **G.** Quadrant plot of DAR versus DEG for the Valproate>14 days (Group C) versus Controls (Group A) comparisons. Volcano plots showing top 20 **H.** DEG and **I.** Differential TF activation in Cardiomyocytes for Valproate>14 days (Group A) versus Controls (Group A) comparisons. **J.** Quadrant plot of DEG from the Group B versus A comparison and the Group C. **K**. KEGG Pathway enrichment of DEG upregulated only in Group B. **L.** KEGG pathway enrichment for DEG only upregulated in Groups A and C.

snRNAseq analyses demonstrated a net reduction in DEG (9 up versus 33 down, **Figure 3B, eTable 8**) and TF activity (**Figure 3C**). GO and KEGG analyses for DEG and differential TF activity were not informative with low numbers of genes enriched per pathway (**eTable 9-13**). Analyses of individual DEG and TF activity (**Figure 3E** and **3F, eTables 8** and **11**) indicated suppression of TF that maintain the contractile cardiomyocyte state (MEF2A, NFATc1) with loss of sarcomere gene programmes (TTN, CMYA5, PALLD), TF suppression of Nrf2-dependent antioxidant defence mechanisms (MAFK, MAFF), and TF activation of a non-proliferative, pro-inflammatory senescent type state (TP53, E2F4, TFDP1, STAT1). Other changes (ELK1, STAT1, STAT6, CCN1, MALAT1, RPL41, NMD3) were typical of stress-response transcription, whereas activation of SIX2, PDX1, EBF1, IRF4, TFAP4 were suggestive of ectopic lineage-inappropriate transcription factor activity.

*For Valproate>14 days versus Controls,* snATACseq demonstrated a net decrease in accessibility to DAR, (12 up versus 15 down). GO and KEGG pathway analyses indicated low levels of enrichment (**eTable 6-7**). Quadrant plots showed complete discordance between DAR and DEG (**Figure 3G, eTable 14-16**).

snRNAseq analyses of individual DEG (no TF passed FDR, **Figure 3H** and **3I**) were consistent with consolidated changes in the cardiomyocyte phenotype with myocardial biogenesis (MT-CO1, MT-CYB, COX8A), cytoskeletal remodelling (DES up, SYNE1 down, COL21A1 down), a nuclear mechanotransduction deficit (SYNE1 down), loss of AIG1 (eliminating Pirh2-mediated p53 ubiquitination and promoting ferroptosis), and suppression of p53 activation (YPEL down) (**Figure 3H** and **3I, eTables 8** and **11**).

A quadrant plot of DEG from the Valproate≤14 days versus Controls, and the Valproate>14 days versus Controls comparisons identified 8 significant DEG associated with changes in myocardial injury (**Figure 3J**). Of these, CEBPB up, MALAT1 up, S100A10 up, RPL41 up, BCKDHB down are consistent with hormetic preconditioning and suppression of ferroptosis (**Figure 3K** and **3L, eTables 14** and **15)** with Valproate≤14 days.

#### Arterial Endothelial Cells

*For Valproate*≤*14 days versus Controls,* snATACseq demonstrated a large net increase in significant DAR (1137 up versus 6 down, **eTable 15-16**). There was complete discordance between DAR and DEG (**Figure 4A**).

**Figure 4.**
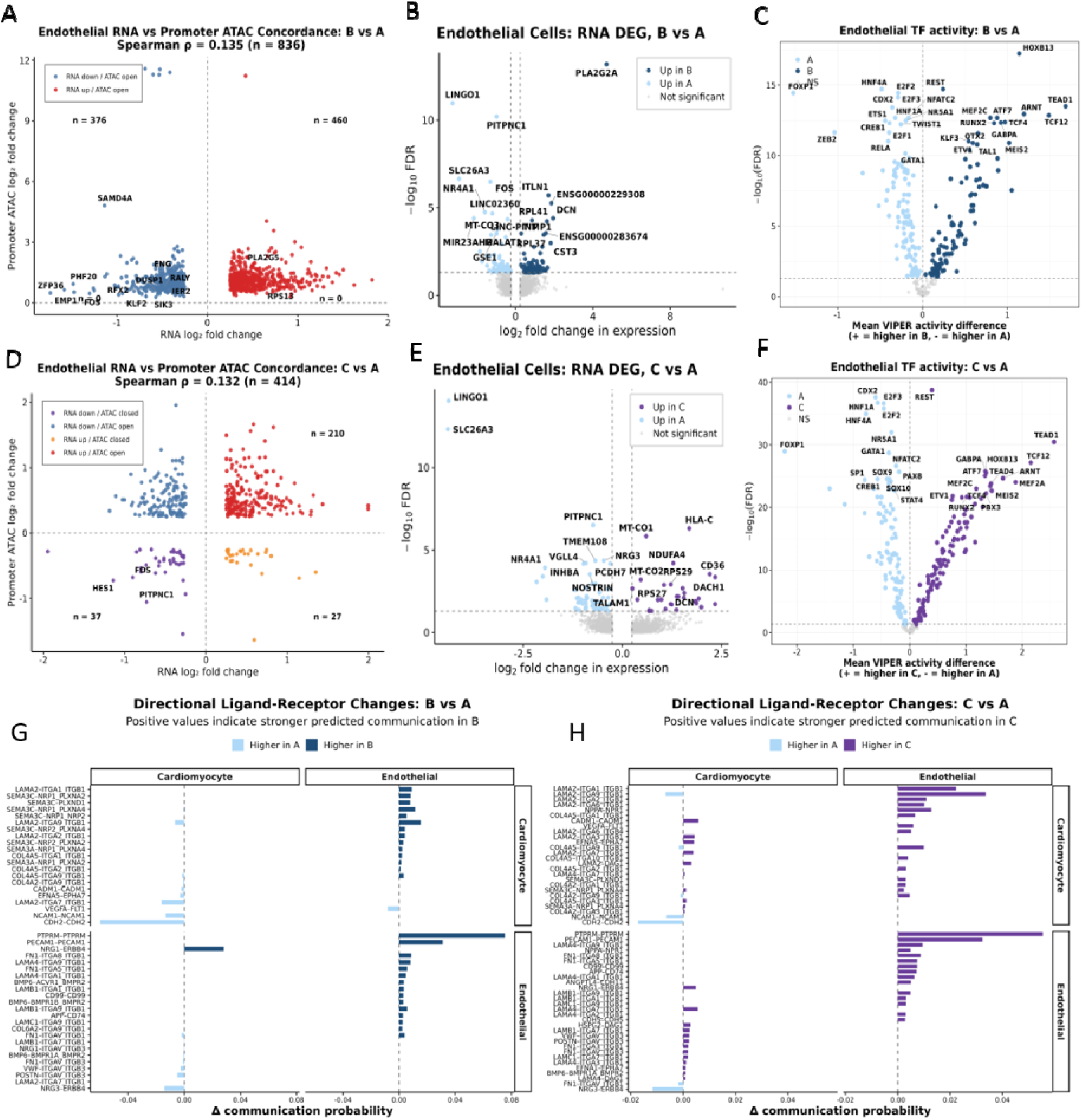
As treated comparison of chromatin accessibility, gene expression, and transcription factor activity in endothelial cells and cell-cell communication analysis. **A.** Quadrant plot of Differential Accessible Regions (DAR) versus Differentially Expressed Genes for the Valproate≤14 days (Group B) versus Controls (Group A) comparisons. Volcano plots showing top 20 **B.** DEG and **C.** Differential TF activation in endothelial cells for Valproate≤14 days (Group B) versus Controls (Group A) comparisons. **D.** Quadrant plot of DAR versus DEG for the Valproate>14 days (Group C) versus Controls (Group A) comparisons. Volcano plots showing top 20 **E.** DEG and **F.** Differential TF activation in endothelial cells for Valproate>14 days (Group A) versus Controls (Group A) comparisons. Directional ligand receptor changes between cardiomyocytes and endothelial cells in **G.** the Valproate14 days (Group B) versus Controls (Group A) comparison, and **H.** Valproate>14 days (Group C) versus Controls (Group A) comparison. Proposed models of **I.** Myocardial resilience and **J.** Myocardial susceptibility to the metabolic stress of surgery associated with Valproate≤14 days (Group B) and Valproate>14 days (Group A) respectively, based on the integrated informatics analyses.

snRNAseq analyses demonstrated a net increase in significant DEG (77 up versus 62 down) and TF activation indicative of activation of stress responses (NRF2, RPL41, RPL37, ATF, CREB1 down), inflammatory lipid signalling (PLA2G2A, ITLN1), and repression of canonical endothelial signalling (ETS1, FOXP1, CREB1, NFATC2), immediate/early proliferative/ angiogenic genes (FOS, NR4A1) and ECM stabilisation (DCN, TIMP1, CST3). Both regulon (TEAD1, RUNX2, ATF7, HOXB13) and DEG analyses (TCF4, TCF12, E2F2/3 down) suggested mesenchymal/ stress adaptive programming characteristic of EndMT phenotypic switching. (**Figure 4B and 4C, eTables 17-23)**.

*For Valproate>14 days versus Controls*, snATACseq demonstrated a small net decrease for DAR (120 up versus 153 down). The DAR versus DEG quadrant plot for Valproate>14 days versus Controls indicated a modest correlation (Spearman rho=0.132) between accessible promoter regions and gene expression (**Figure 4D**) however the numbers of loci with both significant DAR and significant DEG after FDR was low (n=3) and GO and KEGG analyses of concordant DAR and DEG demonstrated low levels of pathway enrichment.

snRNAseq analyses showed signalling consistent with a stable (Hippo/ TEAD, E2F2/3 down) mesenchymal phenotype (TF: ETS1, FOXP1, NFATC2 down, DEG; TALAM1, PCDH7, VGLL4 down), TGFβ signalling (DACH1, DCM), ongoing cell stress (TF; ATF7 up, CREB1 down, DEG; RPS29, RPS27), mitochondrial biogenesis (MT-CO1, MT-CO2, NDUFA4) and loss of eNOS trafficking (NOSTRIN) and normal haemodynamic responsiveness (NR4A1, PITPNC1) (**Figure 4E** and **4F**, **eTables 19-23**).

#### Cell-Cell Communication Analyses

*Valproate*≤*14 days versus Controls* analyses demonstrated loss of cardiomyocyte-cardiomyocyte CDH2-CDH2, NCAM1-MCAM1, CAMD1-CAMD1 EPHA/EFNA5-EPHA7 and endothelial-cardiomyocyte POSTN-ITGAV/ITGB3 signals that maintain contractile cardiomyocyte identity and function. There was increased endothelial-cardiomyocyte SEMA3C signalling that is associated with TP53 senescence programming in cardiomyocytes, as well as increased NRG1-ERBB4 signalling (which activates PI3K/Akt/GSK-3β Ser9) and reduced BMP6-BMPR1A/BMPR2 signalling, all pathways associated with suppression of ferroptosis (**Figure 4G**, and **eTable 23**).

*Valproate>14 days versus Controls*, showed increased NRP1 cardiomyocyte-endothelial-endothelial signalling consistent with natriuretic peptide (ANP, BNP) activation Increased cardiomyocyte-endothelial laminin and collagen signalling, were characteristic of established EndMT. These changes are consistent with a heart failure phenotype (**Figure 4H** and **eTable 23**).

### In vitro experiments

The results of *in vitro* experiments are shown in **Figure 5**. In pre-stressed; IL-6 + H_₂_O_₂_ treated, AC16 cardiomyocytes, short-term exposure to sodium valproate attenuated lipid peroxidation, cell death, and ferroptosis in response to hypoxia reoxygenation. Valproate pre-treatment increased AKT, GSK3β, and p53 gene expression, increased p53 K382 acetylation, and increased GSK3β Ser9 phosphorylation in pre-stressed AC16 cells.

**Figure 5.**
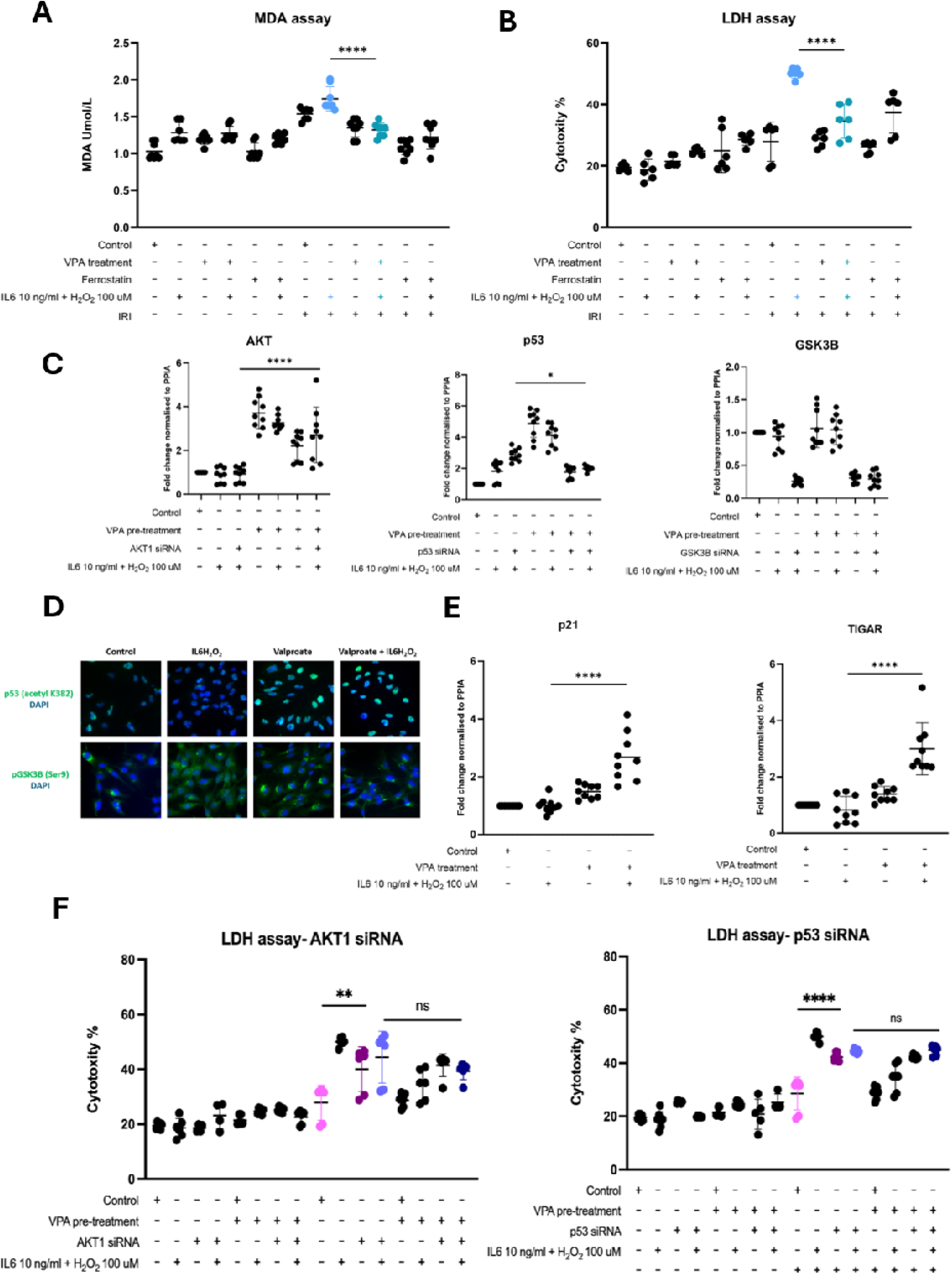
Results of In vitro experiments. See main text. *P<0.05 ****P<0.0001 Statistical tests were one way ANOVA with Bonferroni’s multiple comparisons test. VPA, Valproate. IL6, Interleukin 6. H_₂_O₂, Hydrogen Peroxide. MDA, Malondialdehyde. LDH, Lactate Dehydrogenase. IRI, Ischaemia Reperfusion Injury. AKT, AKT Serine/Threonine Kinase. GSK3β, Glycogen Synthase Kinase 3 Beta. TIGAR, TP53 Induced Glycolysis Regulatory Phosphatase.

Valproate increased expression of p53 anti-apoptotic genes p21 and TIGAR with no increase in expression of the pro apoptotic genes BAX or PUMA (not shown). siRNA silencing of AKT and p53 attenuated the protective effects of valproate in response to IRI.

## Discussion

### Main findings

In an open label dose finding randomised trial, sodium valproate 15mg/kg per day for up to 14 days had higher levels of adherence and low levels of toxicity assessed using CTCAE criteria and serial assessments of blood counts and liver function when compared to treatment >14 days.

Due to high levels of non-adherence across all groups (38%) an as-treated analysis was performed for the mechanism sub-study. Here, participants exposed to IRI and treated with sodium valproate≤14 days demonstrated low levels of myocardial injury relative to controls and Valproate>14 days groups.

snATACseq analyses of myocardial biopsies collected from trial participants at surgery demonstrated significant increases in chromatin accessibility in the Valprotate≤14 days group across all cell types. However, gene expression as demonstrated by integrated snRNAseq demonstrated completely discordant changes, indicating that the increases in accessibility were unprogrammed and not causal for the observed changes in gene expression.

snRNAseq showed that Valproate≤14 days was associated with loss of the contractile phenotype in cardiomyocytes with transcriptional activation indicative of cell distress, hormetic p53 activation, and Akt-GSK3β activation, processes associated with reduced susceptibility to cell death. In vitro experiments showed that these effects were attributable to p53 hyperacetylation, a recognised non-HDACi effect of valproate.(28) These processes were reinforced by changes in endothelial transcription promoting an EndoMT phenotype and endothelial-cardiomyocyte signaling that is also associated with suppression of cell death.

In contrast, Valproate>14 days pre-surgery resulted in signaling changes typical of heart failure, with increased natriuretic peptide signaling, suppression of pathways that confer protection against ferroptosis (AIG, p53), mitochondrial biogenesis, and established EndoMT associated with dysregulated eNOS signaling and loss of normal haemodynamic responsiveness.

Treatment effects of valproate were unrelated to HDACi changes in chromatin accessibility and consistent with hormetic p53 and Akt-GSK-3β activation of cell stress response genes, corroborated by *in vitro* experiments. In contrast, longer duration (>14 days) of treatment results in a heart failure phenotype and increased susceptibility to myocardial injury.

### Clinical Importance

There are no clinically effective myocardial protection strategies for patients undergoing cardiac surgery. Myocardial injury complicates 25% of cardiac surgery procedures where it is the primary cause of death.(1) Further to a previous analyses demonstrating an organ protective of valproate in experimental models of myocardial injury (7) we have now shown that pre-surgery administration of Valproate 15mg/kg/day is well tolerated with low levels of toxicity for up to 14 days. In an as-treated analyses, we showed reductions in myocardial injury, as defined by Devereaux and colleagues, (10) post cardiac surgery with cardioplegic arrest (IRI), and a trend towards reductions in troponin release in the Valproate≤14days treated group. These results are not proof of myocardial protection. The study was designed to test feasibility and safety. However, they are consistent with the results of the integrated snATACseq and snRNAseq analyses of myocardial biopsies collected from trial participants, as well as subsequent mechanistic analyses. Specifically, Valproate≤14days was associated with acute cell stress and activation of cell protection signaling networks that reduce the risks of ferroptosis, an important contributor to post cardiac surgery myocardial injury.(29) In contrast, Valproate>14days was associated with a heart failure like phenotype that is known to increase myocardial susceptibility to ferroptosis.(30) Our interpretation of the combined ITT and as-treated analyses are that valproate 15mg/kg/day ≤14-days showed a positive efficacy signal with low toxicity. This is important in terms of the feasibility of implementation, as almost 50% of cardiac surgery cases are categorised as urgent and require a 5-7 day in-patient wait before surgery, when valproate could be administered.

### Strengths and limitations

Strengths include first, to our knowledge this is the first clinical trial to successfully evaluate the myocardial protection effects of sodium valproate or any HDACi in humans. Second, the ITT analysis did not demonstrate a treatment effect, and the as-treated analysis will have introduced significant bias, given our knowledge of the ITT results. In mitigation, using an ITT approach, when 38% of participants did not receive their allocated treatment for the mechanistic analysis, would have introduced greater bias. The value of this approach was demonstrated by consistent findings across the as-treated analysis of troponin release and myocardial injury, high precision transcriptomics, and *in vitro* experiments. Finally, the trial was terminated after completion of Phase 1 due to the exhaustion of available funds caused by the COVID19 pandemic. These considerations notwithstanding, we suggest that progression to an efficacy trial is warranted based on these proof-of-concept results.

## Conclusions

Pre-surgery administration of sodium valproate is safe and well tolerated in cardiac surgery patients. The direction of the treatment effect on myocardial injury in vivo, defined by serial serum troponin measures post-surgery, in an as treated analysis was consistent with a myocardial protection effect although the study was not designed or powered to detect this. Integrated single cell transcriptomics suggest that contrary to the mechanistic hypothesis, the treatment effects of short-term valproate (≤14 days) were not attributable to increased chromatin accessibility. Rather, the myocardial protection effects were likely attributable to p53 hyperacetylation. Longer duration treatment likely increased myocardial injury susceptibility via epigenetic effects.

## Data Availability

All data produced in the present study are available upon reasonable request to the authors.

https://doi.org/10.5281/zenodo.11091198

## Data Availability

snRNAseq data is available at https://doi.org/10.5281/zenodo.11091198.

Anonymised clinical and biomarker data from the VALCARD study will be made available for ethically approved research with agreement from the study sponsor.

### Code Availability

All bioinformatic analysis were carried out in Unix, Python and R. The codes used for our analyses are available at https://doi.org/10.5281/zenodo.11091198.

## Author Contributions

MR was the principal investigator of the VALCARD trial. NB and CS performed the informatic analyses. SL performed the sequencing and in vitro experiments. LJD, WL, CB, FL, HM, and HA undertook the clinical trial, performed the statistical analyses, and provided the tissue and blood samples for laboratory analyses. GC, TW, VC, and MW co-designed the study and coordinated and supervised laboratory analyses. GJM conceived the study, obtained funding, drafted the manuscript, and is the guarantor of the study. All co-authors have critically reviewed the manuscript and agree to its publication.

## Acknowledgements

The authors would like to express gratitude to the study participants, and clinicians who supported the trial.

